# Short Report. Mental Health-related School Provision and Hospital Contacts among Pupils enrolled in Alternative Provision

**DOI:** 10.64898/2026.11.08.26367536

**Authors:** Sorcha Ní Chobhthaigh, Twyla Greenway-Bailey, Josephine Musanu, Camille Cox, Cyra Neave, Mina Mawi, Amie Buhari, Mel Green, Rochelle A. Burgess, Delan Devakumar, Matthew A. Jay

## Abstract

**Background:** Pupils enrolled in Alternative Provision (AP) represent a highly vulnerable population, with heightened mental health need at risk of long-term educational disengagement. Examining AP enrolment in relation to mental health-related school support and hospital contacts may inform early intervention and prevention approaches to address unmet mental health need.

**Methods:** We analysed population-level linked school and hospital administrative data for 1.7m pupils (born 2000-2002) who attended state schools in England. We examined differences by racial-ethnic group and gender in rates and timing of AP enrolment and recording patterns of mental health-related provision and hospital contacts among pupils with AP enrolment.

**Results:** A majority (62.4%) of pupils with AP enrolment had recorded mental health need, highest among Mixed White-Black Caribbean pupils (74.0%). Among those in AP, 52.0% of girls with a mental health-related hospital contact had school provision, compared to 67.3% boys. For some racial-ethnic groups, schools appeared less likely to record need evident in hospital records; 17.5% boys and 36.2% girls from Mixed White-Asian backgrounds enrolled in AP had mental health-related hospital contact without school provision.

**Conclusion:** Among pupils enrolled in AP, documented mental health need is high but unevenly recorded across systems, with inequalities by racial-ethnic group and gender.

## Introduction

Placements in Alternative Provision (AP) – educational provision outside of mainstream or special schools (1) - have increased dramatically over the past decade (2,3). The majority of pupils enrolled in AP are documented with Special Educational Needs and Disability (SEND) (4), with referral reasons being dominated by concerns related to emotional and behavioural difficulties (5). However, qualitative reports suggest documented figures likely underestimate the extent of placements due to significant mental health need and emphasise without access to appropriate supports in AP complex needs may remain unmet (6,7). Inequalities in access to appropriate mental health supports and use of exclusions may compound the likelihood of AP referrals and under-supported need for some pupils (8).

Recognising pupils referred to AP are particularly vulnerable, we aimed to explore how their mental health needs are captured by school and hospital systems, through SEND for Social, Emotional and Mental Health (SEMH) needs and mental health-related hospital contacts.

## Methods

We analysed the Education and Child Health Insights from Linked Data (ECHILD) database, which contains health records from Hospital Episode Statistics (HES), including information on National Health Service-funded inpatient hospital admissions and outpatient appointments, as well as information about pupils in state-maintained schools and other state-maintained educational settings from the National Pupil Database (NPD). We included all (on-roll) pupils born 2000-2002 who attended state-school in England at any point during mandatory school years (age 5-16) (For more details see: (9)).

We examined outcomes at the intersection of racialisation and gender, using the NPD recording of “ethnicity” as a proxy for racialisation. Due to small numbers, we merged ‘Asian Other & Chinese’ groups (Supplementary Table 1). Pupils without “ethnicity” data (n=9,799) were excluded. Gender was recorded in NPD as male or female, representing assigned “sex”.

NPD recording of AP enrolment indicates a pupil was enrolled in alternative provision or a pupil referral unit. ‘SEND-for-SEMH’ refers to any NPD record of SEND for Behaviour, Emotional & Social Difficulties up to 2013/2014 and Social, Emotional and Mental Health (SEMH) difficulties from 2014/2015 onwards. HES mental health-related hospital contacts include out-patient appointments identified using Mental Health or Psychiatric Team treatment specialty and in-patient admissions with a primary diagnosis related to internalising, externalising, or thought disorder problems or if self-harm was recorded in any diagnostic position.

We calculated rates of AP enrolment, median age-at-first recording using an approximate date of birth derived from NPD, and among pupils with AP enrolment, the proportion recorded with SEND-for-SEMH only, mental health-related contacts only or both.

All data cleaning, preparation and analyses were completed with R software, version 4.4.4 (10) in the Office for National Statistics Secure Research Service.

## Results

### Cohort

Our cohort included 1,696,198 pupils (Supplementary: Figure 1 & Table 2).

### Rates

Rates of AP enrolment differed significantly at the intersection of racial-ethnic groups and gender (Supplementary: Figure 2 & Table 3). Rates of AP enrolment were higher among pupils with SEND-for-SEMH (5.3 times higher boys; 7.5 times higher girls) and mental health-related hospital contact (5.8 times higher boys; 5.3 times higher girls).

### Age

For most groups, median age-at-first AP enrolment clustered at 13-14 years for boys, younger for White Irish and Irish Traveller boys age 12, and 14-15 for girls, again youngest for Irish Traveller girls (Supplementary Table 4). For pupils enrolled in AP, across groups the median age-at-first SEND-for-SEMH was 8.3 for boys and 11.2 for girls, and mental health-related hospital contact was 11.1 for boys and 13.4 for girls.

### Recording Patterns

Among pupils with AP enrolment, boys are more often recorded by school-only (SEND-for-SEMH only 32.4% boys, 15.7% girls) and girls by hospital services only (Hospital-only 18.5% girls, boys 10.8%) (Table 1). Girls also more often had neither recording (Neither 45.9%, boys 34.6%). Of pupils with a mental health-related hospital contact, 67.3% of boys were also recorded with SEND-for-SEMH, compared with 52.0% of girls. Recording patterns varied considerably across racial-ethnic groups; where data was available, hospital-only recordings were highest among Mixed White-Asian girls (36.2%) and boys (17.5%), while dual recordings were highest among Mixed White-Black Caribbean boys (28.0%) and girls (31.6%).

**Table 1.** Recording Patterns: Proportions of (a) Neither SEND-for-SEMH or Mental Health-related Contact (b) SEND-for-SEMH-only (c) Mental Health-related Contact-only (d) Both Among Pupils Ever Recorded with AP Enrolment. In table ‘x’ used to denote suppression due to small cell counts.

|  | **White British** | **White Irish** | **White Other** | **Mixed White-Black African** | **Mixed White-Black Caribbean** | **Mixed White-Asian** | **Mixed Other** | **Black African** | **Black Caribbean** | **Black Other** | **Indian** | **Pakistani** | **Bangladeshi** | **Asian Other & Chinese** | **Romani** | **Irish Traveller** | **Other** | **Combined** |
|---|---|---|---|---|---|---|---|---|---|---|---|---|---|---|---|---|---|---|
| **Neither School nor Hospital Mental Health-related Recording (%)** |  |  |  |  |  |  |  |  |  |  |  |  |  |  |  |  |  |  |
| **All** | 30.9 | 40.5 | 64.2 | 39.3 | 26.0 | 35.3 | 38.9 | 55.7 | 29.7 | 36.3 | 76.7 | 58.9 | 62.7 | 72.6 | 76.6 | 64.7 | 70.7 | 37.6 |
| **Boys** | 28.6 | 37.7 | 61.6 | 37.1 | 22.3 | 35.0 | 40.0 | 50.6 | 27.2 | 33.8 | 70.4 | 54.9 | 63.1 | 74.6 | 69.3 | 59.7 | 66.8 | 34.6 |
| **Girls** | 37.8 | 47.8 | 69.4 | 45.7 | 36.0 | 36.2 | 36.1 | 69.0 | 40.0 | 44.2 | 86.8 | 70.4 | 61.8 | 68.3 | 86.7 | 73.8 | 81.0 | 45.9 |
| **School SEND-for-SEMH-only (%)** |  |  |  |  |  |  |  |  |  |  |  |  |  |  |  |  |  |  |
| **All** | 30.4 | 21.4 | 16.6 | 28.6 | 35.6 | 19.3 | 26.2 | 23.6 | 42.8 | 35.7 | x | 18.6 | x | 7.2 | 15.1 | 18.5 | 8.2 | 27.9 |
| **Boys** | 34.8 | x | 20.3 | x | 41.7 | x | 29.3 | 27.7 | 47.0 | x | x | x | x | x | x | x | x | 32.4 |
| **Girls** | 17.8 | x | 9.4 | x | 19.1 | x | 18.5 | 13.1 | 25.5 | x | x | x | x | x | x | x | x | 15.7 |
| **Mental Health-related Hospital Contact-only (%)** |  |  |  |  |  |  |  |  |  |  |  |  |  |  |  |  |  |  |
| **All** | 13.7 | 13.1 | 10.4 | 12.1 | 9.4 | 23.3 | 13.9 | 12.0 | 5.5 | 12.1 | 13.1 | 11.8 | 14.4 | 14.4 | x | x | 13.1 | 12.9 |
| **Boys** | 11.1 | x | 9.4 | x | 8.1 | 17.5 | 9.7 | 12.7 | 4.2 | x | x | 11.8 | x | 12.7 | x | x | 15.1 | 10.8 |
| **Girls** | 21.1 | x | 12.3 | x | 13.2 | 36.2 | 24.4 | 10.1 | 10.9 | x | x | 12.0 | x | 18.3 | x | x | 7.9 | 18.5 |
| **Both School & Hospital (%)** |  |  |  |  |  |  |  |  |  |  |  |  |  |  |  |  |  |  |
| **All** | 25.0 | 25.0 | 8.8 | 20.0 | 28.9 | 22.0 | 21.0 | 8.7 | 22.0 | 15.9 | x | 10.6 | x | 5.7 | x | x | 8.0 | 21.6 |
| **Boys** | 25.5 | x | 8.8 | x | 28.0 | x | 21.0 | 9.1 | 21.6 | x | x | x | x | x | x | x | x | 22.2 |
| **Girls** | 23.4 | x | 9.0 | x | 31.6 | x | 21.0 | 7.7 | 23.6 | x | x | x | x | x | x | x | x | 20.0 |

## Discussion

### What is already known on this topic

Racial-ethnic and gender inequities exist in school responses to mental health need, including SEND-for-SEMH identification, use of exclusions and off-rolling (8,11), increasing the likelihood of inequities in educational pathways and AP placement referrals. Systematic differences in referrals and pathways to appropriate mental health care through the health system further influence the likelihood of unmet need across groups (12–14).

### What this study adds

Just under two-thirds of boys and over half of girls with AP enrolment have recorded mental health needs in school, hospital or both. System recording of need varies significantly across genders and racial-ethnic groups. The young age of school and hospital recording of mental health need among these pupils suggests that, for some, vulnerability may be identified from a young age. The clustering of AP enrolment 13-15 years suggests for those with identified need supports in place were insufficient or for those without there was a missed opportunity to prevent need escalation, particularly following the transition to secondary school. Among pupils ever recorded with AP enrolment, the majority of boys who had a mental health-related hospital contact were also recorded with SEND-for-SEMH, yet approximately half of girls with a mental health-related hospital contact had dual recording, suggesting the distress of girls with AP enrolment is less likely to be formally identified in the school system. For some racial-ethnic groups, this appears amplified, such as for Mixed White-Asian pupils with the highest proportion of hospital-only recordings meaning need that surfaced in the hospital system was not formally identified in school.

### Limitations

HES data contains in-patient and out-patient hospital records, including Child and Adolescent Mental Health Services (CAMHS) operating at hospital sites, but not community-based CAMHS. The AP census began 2008/2009 as such AP enrolment is likely underestimated, and the Pupil Referral Unit census was only available 2010-2013. NPD racial-ethnic categories inevitably invisibilise some groups and small numbers prevent reporting of specific recording patterns for some groups. As AP includes hospital-based educational provision for children receiving in-patient care, and girls are disproportionality admitted for stress-related presentations (15), future research should carefully examine inequities in the transition back to school following discharge.

## Conclusion

A majority of pupils ever enrolled in AP had documented mental health needs, however, these were inconsistently captured across systems based on racial-ethnic group and gender. Many pupils experiencing needs that warranted access to hospital-based mental health services were not recorded with SEND-for-SEMH, suggesting gaps between systems and discrimination in access to appropriate supports.

To reduce risk of escalating unmet need and referrals to AP, mainstream schools must be better equipped to create inclusive educational environments, responsive to early indicators of pupils emotional and behavioural needs. Where specialist provision is in the best interest of the pupil, a standard of evidence-based and trauma-informed practice must be established nationally.

## Ethics

This study uses linked, de-identified data from Hospital Episode Statistics and the National Public Database as part of ECHILD. Ethical approval for the ECHILD project was granted by the National Research Ethics Service (17/LO/1494), NHS Health Research Authority Research Ethics Committee (20/EE/0180 and 21/SW/0159) and is overseen by the UCL Great Ormond Street Institute of Child Health’s Joint Research and Development Office (20PE16). Consent from patients is not required. Study-specific ethical approval was obtained from UCL Research Ethics Committee (24643/001).

## Data Availability

The ECHILD Database uses data from the Department for Education (DfE) and data provided by patients and collected by the National Health Service as part of their care and support. Source data can be accessed by researchers by applying directly to NHS England or DfE. ECHILD Data may be obtained from a third party and is not publicly available, access can be applied for through the ECHILD team via the website: [https://www.echild.ac.uk/]

## Funding Statement

Sorcha Ní Chobhthaigh is funded by the Medical Research Council (MRC) grant MR/N013867/1. ECHILD is supported by ADR UK (Administrative Data Research UK), an Economic and Social Research Council (part of UK Research and Innovation) programme.

## Conflict of interest statement

There are no conflicts of interest to disclose.

## Author Contributions

SNC is the guarantor: SNC had access to the data in the study and takes responsibility for the integrity of the data and the accuracy of the data analysis. SNC conceptualised and designed the study, performed the data analyses, and wrote the initial draft manuscript. CC, JM and TGB contributed to study design, analysis decisions, interpretation of results and co-development of recommendations. CN, MG, MM and AB contributed to interpretation of results and co-development of recommendations. MJ supported data management, code development and supervised the analytical approach. RB supervised the participatory methodology. DD provided supervisory support throughout. All authors reviewed, commented, and approved the final manuscript.

## Acknowledgements

We would like to express our appreciation to additional and former stakeholder advisors (in addition to those who are named authors). Their meaningful contributions and thoughtful insights shaped this project, contextualisation and co-developed recommendations. We also extend our wholehearted gratitude to the No More Exclusions coalition for their valuable feedback and insights.

We thank all the children, young people, parents and carers who contributed to the ECHILD project. We gratefully acknowledge all children and families whose de-identified data are used in this research.

The ECHILD Database uses data from the Department for Education (DfE) and data provided by patients and collected by the National Health Service as part of their care and support. Source data can be accessed by researchers by applying directly to NHS England or DfE. ECHILD Data may be obtained from a third party and is not publicly available, access can be applied for through the ECHILD team via https://www.echild.ac.uk/. We are grateful to the Office for National Statistics (ONS) for providing the trusted research environment for the ECHILD Database. ONS agrees that the results in the attached document may be published. This does not imply ONS’ acceptance of the validity of the methods used to btain these figures, or of any analysis of the results. The DfE, NHSE and ONS do not accept responsibility for any inferences or conclusions derived by the authors. The views in this publication do not necessarily reflect the views of UCL. We acknowledge the contribution of the wider ECHILD Database support and programme management.

## Supplementary Files

- Supplementary Table 1: Racial-Ethnic Groups
- Supplementary Figure 1: Cohort Flow-chart
- Supplementary Table 2: Cohort Overview with Median Age at School Enrolment
- Supplementary Table 3: Rates per 1,000 children and young people
- Figure 1. Rates of AP enrolment per 1,000 pupils (a) All pupils (b) Among pupils ever recorded with SEND-for-SEMH (c) Among pupils ever recorded with Mental Health-related Hospital Contact
- Supplementary Table 4: Median Age-at-First

## Notes

### Competing Interest Statement

The authors have declared no competing interest.

