## Supplementary Files for "Short Report. Mental Health-related School Provision and Hospital Contacts among Pupils enrolled in Alternative Provision"

#### Supplementary Table 1: Racial-Ethnic Groups

| *National Pupil Database 'Ethnic Minor' variable categories* | *Study racial-ethnic categories* |
|---|---|
| White British | White British |
| White Irish | White Irish |
| White Other | White Other |
| Mixed White-Black African | Mixed White-Black African |
| Mixed White-Black Caribbean | Mixed White-Black Caribbean |
| Mixed White-Asian | Mixed White-Asian |
| Mixed Other | Mixed Other |
| Black African | Black African |
| Black Caribbean | Black Caribbean |
| Black Other | Black Other |
| Indian | Indian |
| Pakistani | Pakistani |
| Bangladeshi | Bangladeshi |
| Asian Other | Asian Other & Chinese |
| Chinese | Romani |
| Romani | Irish Traveller |
| Irish Traveller | Other |
| Other | |

#### Supplementary Figure 1: Cohort Flow-chart

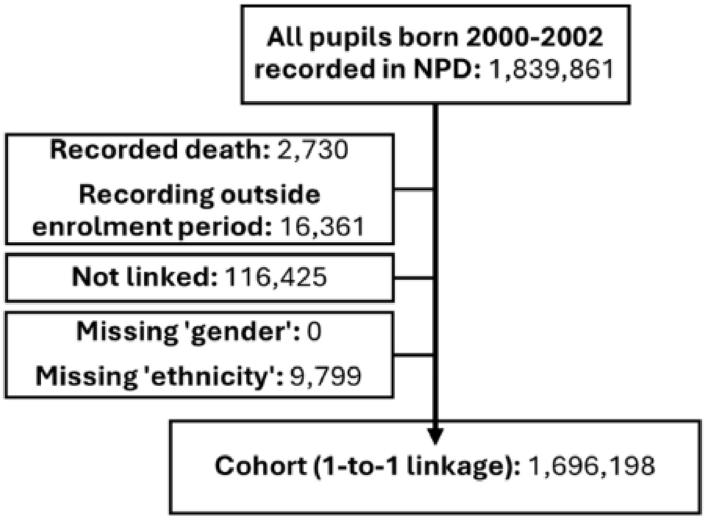

### Supplementary Table 2: Cohort Overview with Median Age at School Enrolment

| | White British | White Irish | White Other | Mixed White-Black African | Mixed White-Black Caribbean | Mixed White-Asian | Mixed Other | Black African | Black Caribbean | Black Other | Indian | Pakistani | Bangladeshi | Asian Other & Chinese | Romani | Irish Traveller | Other | Combined |
|---|---|---|---|---|---|---|---|---|---|---|---|---|---|---|---|---|---|---|
| **Pupils Included in the Study *n (%)*** | | | | | | | | | | | | | | | | | | |
| **All** *(row%)* | 1209077 *(71.3)* | 5882 *(0.3)* | 97499 *(5.7)* | 8733 *(0.5)* | 20925 *(1.2)* | 16522 *(1.0)* | 27260 *(1.6)* | 58943 *(3.5)* | 20902 *(1.2)* | 10495 *(0.6)* | 46993 *(2.8)* | 70116 *(4.1)* | 27407 *(1.6)* | 35350 *(2.1)* | 6407 *(0.4)* | 1681 *(0.1)* | 32006 *(1.9)* | 1696198 *(100)* |
| **Male** *(col%)* | 623973 *(51.6)* | 3028 *(51.5)* | 50044 *(51.3)* | 4455 *(51.0)* | 10880 *(52.0)* | 8498 *(51.4)* | 14146 *(51.9)* | 30259 *(51.3)* | 10973 *(52.5)* | 5408 *(51.5)* | 24540 *(52.2)* | 36431 *(52.0)* | 14088 *(51.4)* | 18434 *(52.1)* | 3305 *(51.6)* | 879 *(52.3)* | 16998 *(53.1)* | 876339 *(51.7)* |
| **Female** *(col%)* | 585104 *(48.4)* | 2854 *(48.5)* | 47455 *(48.7)* | 4278 *(49.0)* | 10045 *(48.0)* | 8024 *(48.6)* | 13114 *(48.1)* | 28684 *(48.7)* | 9929 *(47.5)* | 5087 *(48.5)* | 22453 *(47.8)* | 33685 *(48.0)* | 13319 *(48.6)* | 16916 *(47.9)* | 3102 *(48.4)* | 802 *(47.7)* | 15008 *(46.9)* | 819859 *(48.3)* |
| **Median Age at School Enrolment *(years)*** | | | | | | | | | | | | | | | | | | |
| **All** | 5.20 | 5.21 | 5.46 | 5.21 | 5.20 | 5.21 | 5.21 | 5.21 | 5.20 | 5.21 | 5.21 | 5.21 | 5.21 | 5.29 | 6.30 | 5.21 | 5.30 | 5.21 |
| **Male** | 5.20 | 5.21 | 5.38 | 5.21 | 5.20 | 5.21 | 5.21 | 5.21 | 5.20 | 5.21 | 5.21 | 5.21 | 5.21 | 5.29 | 6.21 | 5.21 | 5.30 | 5.21 |
| **Female** | 5.20 | 5.21 | 5.46 | 5.21 | 5.20 | 5.21 | 5.21 | 5.21 | 5.20 | 5.21 | 5.21 | 5.21 | 5.21 | 5.29 | 6.30 | 5.21 | 5.29 | 5.21 |

#### Supplementary Table 3: Rates per 1,000 children and young people

| | White British | White Irish | White Other | Mixed White-Black African | Mixed White-Black Caribbean | Mixed White-Asian | Mixed Other | Black African | Black Caribbean | Black Other | Indian | Pakistani | Bangladeshi | Asian Other & Chinese | Romani | Irish Traveller | Other | Combined |
|---|---|---|---|---|---|---|---|---|---|---|---|---|---|---|---|---|---|---|
| **(a) Alternative Provision** ***Rates per 1,000 (95% CI)*** | | | | | | | | | | | | | | | | | | |
| **All** | 10.3 (10.2-10.5) | 14.3 (11.2-17.3) | 9.5 (8.9-10.1) | 16.0 (13.4-18.7) | 24.3 (22.2-26.4) | 9.1 (7.6-10.5) | 15.0 (13.6-16.4) | 10.3 (9.5-11.1) | 26.9 (24.7-29.1) | 17.3 (14.8-19.8) | 3.7 (3.2-4.3) | 5.9 (5.3-6.5) | 4.3 (3.5-5.1) | 7.4 (6.5-8.3) | 47.4 (42.2-52.7) | 70.8 (58.5-83.1) | 14.1 (12.8-15.4) | 10.6 (10.4-10.7) |
| **Male** | 14.9 (14.6-15.2) | 20.1 (15.1-25.1) | 12.3 (11.4-13.3) | 23.6 (19.1-28.0) | 34.2 (30.8-37.6) | 12.1 (9.8-14.4) | 20.5 (18.2-22.8) | 14.6 (13.2-15.9) | 41.3 (37.6-45.0) | 25.7 (21.5-29.9) | 4.4 (3.6-5.2) | 8.4 (7.5-9.3) | 6.0 (4.7-7.2) | 9.8 (8.4-11.2) | 53.3 (45.6-60.9) | 87.6 (68.9-106.3) | 19.1 (17.1-21.2) | 15.0 (14.7-15.2) |
| **Female** | 5.5 (5.3-5.7) | 8.1 (4.8-11.3) | 6.5 (5.8-7.3) | 8.2 (5.5-10.9) | 13.5 (11.3-15.8) | 5.9 (4.2-7.5) | 9.1 (7.5-10.7) | 5.9 (5.0-6.7) | 11.1 (9.0-13.1) | 8.5 (5.9-11.0) | 3.0 (2.3-3.7) | 3.2 (2.6-3.8) | 2.6 (1.7-3.4) | 4.8 (3.8-5.9) | 41.3 (34.3-48.3) | 52.4 (37.0-67.8) | 8.4 (6.9-9.9) | 5.9 (5.7-6.0) |
| **(b) Alternative Provision among Pupils Ever Recorded with SEND-for-SEMH** ***Rates per 1,000 (95% CI)*** | | | | | | | | | | | | | | | | | | |
| **All** | 70.7 (69.1-72.3) | 92.0 (64.5-119.5) | 51.6 (45.1-58.0) | 79.7 (61.5-97.9) | 103.2 (92.6-113.8) | 57.3 (43.5-71.1) | 82.4 (71.2-93.5) | 45.1 (38.9-51.2) | 98.5 (88.9-108.1) | 88.4 (71.4-105.5) | 16.3 (8.8-23.8) | 36.4 (30.0-42.7) | 21.5 (13.5-29.5) | 37.8 (25.3-50.2) | 96.6 (73.7-119.4) | 136.7 (94.6-178.8) | 44.3 (34.4-54.3) | 69.0 (67.6-70.4) |
| **Male** | 81.8 (79.7-83.8) | x | 57.2 (49.1-65.4) | 89.9 (67.0-112.7) | 118.1 (104.6-131.6) | 65.8 (48.0-83.6) | 90.9 (76.8-104.9) | 51.5 (43.8-59.2) | 118.0 (105.7-130.3) | 102.5 (80.9-124.0) | x | 42.7 (34.6-50.9) | x | 38.7 (23.2-54.2) | 104.7 (77.0-132.4) | x | 50.8 (38.2-63.4) | 79.7 (77.9-81.5) |
| **Female** | 45.1 (42.7-47.4) | x | 39.3 (29.3-49.3) | 55.6 (27.3-83.8) | 70.1 (54.1-86.0) | 38.6 (18.0-59.1) | 63.9 (46.2-81.5) | 28.6 (19.2-37.9) | 50.5 (37.4-63.6) | 53.0 (27.7-78.2) | x | 20.2 (11.2-29.2) | x | 35.9 (15.1-56.8) | 74.7 (35.6-113.8) | x | 28.9 (14.0-43.9) | 44.2 (42.2-46.3) |
| **(c) Alternative Provision among Pupils Ever Recorded with Mental Health-related Hospital Contact** ***Rates per 1,000 (95% CI)*** | | | | | | | | | | | | | | | | | | |
| **All** | 55.9 (54.4-57.4) | 77.1 (51.4-102.8) | 41.0 (35.1-46.8) | 71.9 (51.7-92.1) | 95.1 (82.4-107.8) | 57.4 (44.2-70.7) | 68.6 (57.8-79.5) | 65.6 (54.5-76.7) | 102.2 (87.0-117.5) | 95.0 (70.2-119.8) | 24.5 (16.6-32.5) | 35.2 (28.2-42.2) | 31.2 (20.0-42.4) | 47.2 (34.8-59.7) | 138.1 (87.9-188.4) | 246.9 (153.0-340.8) | 59.9 (48.2-71.6) | 56.6 (55.2-58.0) |
| **Male** | 86.3 (83.5-89.0) | x | 61.7 (50.6-72.8) | 110.7 (74.0-147.5) | 135.9 (114.5-157.3) | 83.5 (59.9-107.1) | 92.2 (74.0-110.5) | 103.0 (83.5-122.5) | 152.7 (127.3-178.2) | 125.9 (86.4-165.5) | x | 54.4 (41.8-67.1) | 47.4 (26.6-68.2) | 60.8 (40.0-81.5) | x | x | 93.2 (73.0-113.4) | 86.8 (84.3-89.3) |
| **Female** | 30.4 (28.9-32.0) | x | 26.1 (19.9-32.3) | 40.5 (19.7-61.2) | 57.3 (43.4-71.3) | 36.5 (22.2-50.9) | 48.3 (35.7-60.8) | 30.4 (19.7-41.1) | 50.7 (35.0-66.4) | 63.7 (34.4-93.0) | x | 18.4 (11.4-25.4) | 18.9 (7.3-30.6) | 35.9 (21.2-50.7) | x | x | 26.5 (15.3-37.7) | 31.2 (29.8-32.6) |

***Figure 1. Rates of AP enrolment per 1,000 pupils (a) All pupils (b) Among pupils ever recorded with SEND-for-SEMH (c) Among pupils ever recorded with Mental Health-related Hospital Contact***

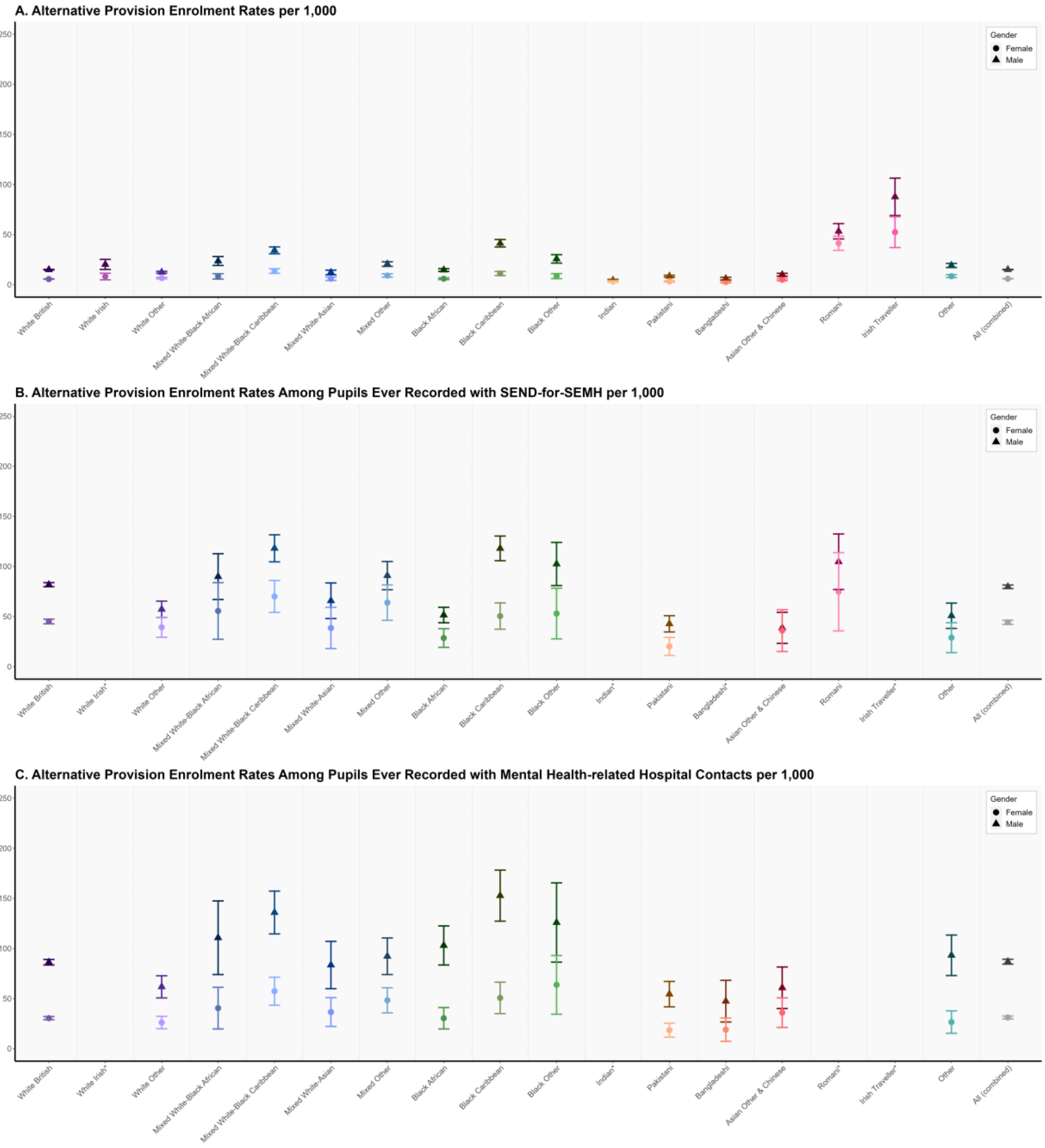

#### Supplementary Table 4: Median Age-at-First

| | White British | White Irish | White Other | Mixed White-Black African | Mixed White-Black Caribbean | Mixed White-Asian | Mixed Other | Black African | Black Caribbean | Black Other | Indian | Pakistani | Bangladeshi | Asian Other & Chinese | Romani | Irish Traveller | Other | Combined |
|---|---|---|---|---|---|---|---|---|---|---|---|---|---|---|---|---|---|---|
| **(a) Age at First Alternative Provision Enrolment: All school-aged children, *Median Age (years)*** | | | | | | | | | | | | | | | | | | |
| **All** | 13.9 | 13.7 | 15.3 | 14.1 | 13.9 | 13.8 | 14.3 | 14.3 | 13.8 | 14.9 | 14.9 | 14.9 | 14.9 | 15.0 | 14.0 | 12.0 | 15.5 | 14.1 |
| **Male** | 13.5 | 12.5 | 15.1 | 13.8 | 13.5 | 13.0 | 14.1 | 13.7 | 13.3 | 14.8 | 14.7 | 14.8 | 14.8 | 15.0 | 14.1 | 12.4 | 15.5 | 13.8 |
| **Female** | 14.8 | 14.6 | 15.5 | 14.7 | 14.9 | 15.0 | 14.7 | 15.2 | 15.0 | 15.4 | 15.1 | 15.1 | 15.2 | 15.0 | 13.9 | 11.9 | 15.6 | 14.9 |
| **(b) Age at First Alternative Provision Enrolment: Pupils ever recorded with SEND-for-SEMH, *Median Age (years)*** | | | | | | | | | | | | | | | | | | |
| **All** | 13.8 | 12.3 | 14.6 | 13.7 | 13.7 | 14.6 | 14.2 | 13.4 | 13.9 | 14.3 | x | 14.8 | x | 13.6 | 13.0 | 12.0 | 14.9 | 13.8 |
| **Male** | 13.4 | x | 14.3 | x | 13.1 | x | 14.1 | 12.6 | 13.5 | x | x | x | x | x | x | x | x | 13.5 |
| **Female** | 14.8 | x | 14.8 | x | 14.9 | x | 14.7 | 14.7 | 15.1 | x | x | x | x | x | x | x | x | 14.8 |
| **(c) Age at First Alternative Provision Enrolment: Pupils ever recorded with Mental Health-related Hospital Contact, *Median Age (years)*** | | | | | | | | | | | | | | | | | | |
| **All** | 14.0 | 13.1 | 14.3 | 14.1 | 13.1 | 14.3 | 14.0 | 14.0 | 13.5 | 14.7 | 13.3 | 14.3 | 14.2 | 13.2 | 14.4 | 13.1 | 14.6 | 14.0 |
| **Male** | 13.3 | x | 13.9 | x | 12.4 | 13.5 | 13.3 | 13.5 | 12.0 | x | x | 14.2 | x | 12.3 | x | x | 14.0 | 13.3 |
| **Female** | 14.9 | x | 14.8 | x | 14.7 | 15.1 | 14.8 | 14.6 | 15.1 | x | x | 15.0 | x | 14.2 | x | x | 15.4 | 14.9 |
| **(d) Age at First SEND-for-SEMH: Pupils ever recorded with Alternative Provision, *Median Age (years)*** | | | | | | | | | | | | | | | | | | |
| **All** | 8.7 | 7.1 | 10.7 | 8.6 | 7.7 | 10.8 | 8.0 | 8.6 | 7.6 | 8.3 | x | 11.1 | x | 9.7 | 11.0 | 9.3 | 10.3 | 8.7 |
| **Male** | 8.3 | x | 9.8 | x | 7.5 | x | 7.7 | 8.1 | 7.4 | x | x | x | x | x | x | x | x | 8.3 |
| **Female** | 11.2 | x | 12.4 | x | 9.1 | x | 11.5 | 10.9 | 10.6 | x | x | x | x | x | x | x | x | 11.2 |
| **(e) Age at First Mental Health-related Hospital Contact: Pupils ever recorded with Alternative Provision, *Median Age (years)*** | | | | | | | | | | | | | | | | | | |
| **All** | 11.7 | 11.2 | 12.9 | 12.2 | 12.4 | 11.7 | 12.2 | 12.4 | 11.8 | 12.9 | 12.3 | 12.5 | 13.1 | 12.1 | 13.5 | 12.0 | 12.7 | 11.9 |
| **Male** | 10.9 | x | 12.3 | x | 11.3 | 10.3 | 11.7 | 11.7 | 10.9 | x | x | 12.3 | x | 12.0 | x | x | 12.1 | 11.1 |
| **Female** | 13.3 | x | 13.6 | x | 13.6 | 13.2 | 13.5 | 13.5 | 13.8 | x | x | 13.0 | x | 12.8 | x | x | 13.6 | 13.4 |
